# Psychological inoculation protects young women in South Africa against HIV prevention misinformation: Evidence from a randomised controlled trial

**DOI:** 10.64898/2026.10.27.26366282

**Authors:** Brendan Maughan-Brown, Divya Pandey, Simamkele Bokolo, Teniola I. Egbe, Linda-Gail Bekker, Candice Chetty-Makkan, Lawrence Long, Sophie Pascoe, Alisa Stevens Shield, Sander van der Linden, Harsha Thirumurthy, Alison M. Buttenheim

**Affiliations:** Southern Africa Labour and Development Research Unit (SALDRU), University of Cape Town, Cape Town, South Africa; Department of Medical Ethics and Health Policy, University of Pennsylvania, Philadelphia; Center for Health Incentives and Behavioral Economics, University of Pennsylvania, Philadelphia; Health Economics and Epidemiology Research Office, Faculty of Health Sciences, University of the Witwatersrand, Johannesburg; Desmond Tutu HIV Centre, University of Cape Town, Cape Town; Department of Global Health, Boston University, Boston; Department of Biostatistics, Epidemiology and Informatics, University of Pennsylvania, Philadelphia; Department of Psychology, University of Cambridge, United Kingdom; Department of Family and Community Health, University of Pennsylvania, Philadelphia

**Author notes:** Corresponding author: Brendan Maughan-Brown.

## Abstract

**Background:** Misinformation decreases public confidence in health technologies and willingness to use them. Psychological inoculation, or “prebunking”, exposes people to misleading claims before they spread widely and refutes the claims and embedded persuasion tactics. Evidence on its effectiveness for HIV-related misinformation, particularly in sub-Saharan Africa and among young women (a population highly vulnerable to HIV), is limited.

**Methods:** We conducted a randomised controlled trial (July to October 2025) among 2,163 women aged 18-29 years recruited online in South Africa. Participants were randomly assigned to view two short TikTok-style inoculation videos addressing common myths about an HIV vaccine, or attention-matched control videos on nutrition and diabetes. All participants then viewed multiple false claims presented as social media posts. The primary outcome was intention to receive an HIV vaccine if available (1-10 scale), assessed immediately post-intervention and at three-week follow-up. Secondary outcomes included likelihood of recommending the vaccine; and perceived credibility, emotional impact, and shareability of misinformation claims. We used linear regression models to estimate intervention effects.

**Results:** Misinformation exposure reduced vaccine intentions by 0.63 points in the control group, while intentions increased by 0.30 points in the intervention group. Controlling for covariates and baseline intentions, post-exposure vaccine intentions were 0.92 points higher in the intervention group (95% CI 0.78, 1.06; p<0.01), a 12% relative increase. Intervention participants were also more likely to recommend the vaccine (+0.74; 95% CI 0.61, 0.88; p<0.01). Inoculation reduced perceived credibility, emotional distress, and sharing intentions across all false claims. Effects persisted at three-week follow-up (vaccine intentions: +0.97; 95% CI 0.78, 1.17; p<0.01).

**Conclusion:** Brief, scalable psychological inoculation delivered via social-media-style videos protected young women’s HIV vaccine intentions, with effects generalising to novel claims and persisting over three weeks. Prebunking campaigns may safeguard public confidence in other new HIV prevention technologies like lenacapavir.

## Introduction

Expanded HIV prevention and treatment services across sub-Saharan Africa over the past 20 years has enabled millions to live longer, healthier lives, yet HIV incidence remains high – particularly among adolescent girls and young women. Many prevention efforts have had only modest success, with low uptake and low sustained use of effective prevention options like oral pre-exposure prophylaxis (PrEP) among young women.^1^ As a result, the development of new prevention options, including an HIV vaccine, remains important for ending the HIV epidemic.

The recent discovery of highly-effective long-acting injectable PrEP represents a major opportunity to accelerate progress toward an HIV-free generation. Among new modalities, lenacapavir is the first twice-yearly injectable PrEP shown to offer substantial protection against HIV acquisition.^2^ Lenacapavir has received swift global support, including World Health Organization prequalification and record-speed approval in South Africa.^3,4^

While ensuring adequate supply of new biomedical innovations like lenacapavir is a prerequisite for altering the trajectory of the HIV epidemic, addressing the barriers to demand for such innovations is equally important. Prior experience with innovations like COVID-19 and human papillomavirus (HPV) vaccines has demonstrated how viral misinformation – especially regarding safety, ingredients, and scientific and policymaker intent – can erode public confidence and significantly undermine uptake.^5–7^ Identifying effective and scalable approaches to countering misinformation about new health products – particularly before it becomes viral and widespread – is thus an important priority for policymakers and public health practitioners.

We investigated psychological inoculation as a strategy to proactively build resistance to HIV prevention misinformation, focusing here on a yet-to-be developed HIV vaccine. Analogous to medical inoculation, psychological inoculation entails exposing individuals to weakened versions of misinformation or misleading arguments, alongside refutations or explanations that highlight the common fallacies and manipulative tactics used in misinformation campaigns.^8,9^ This “prebunking” process aims to build cognitive resistance among individuals before they encounter stronger, more persuasive forms of misinformation – it gets ahead of misinformation before it can do damage.^8^ Figure 1 depicts the theory of psychological inoculation. Prebunking may be more effective than debunking, which counters false claims after they’ve already spread and taken hold, because of the continued influence effect: the psychological phenomenon where outdated or false information continues to influence a person’s thinking and decision-making even after it has been corrected or retracted.^11^

**Figure 1.**
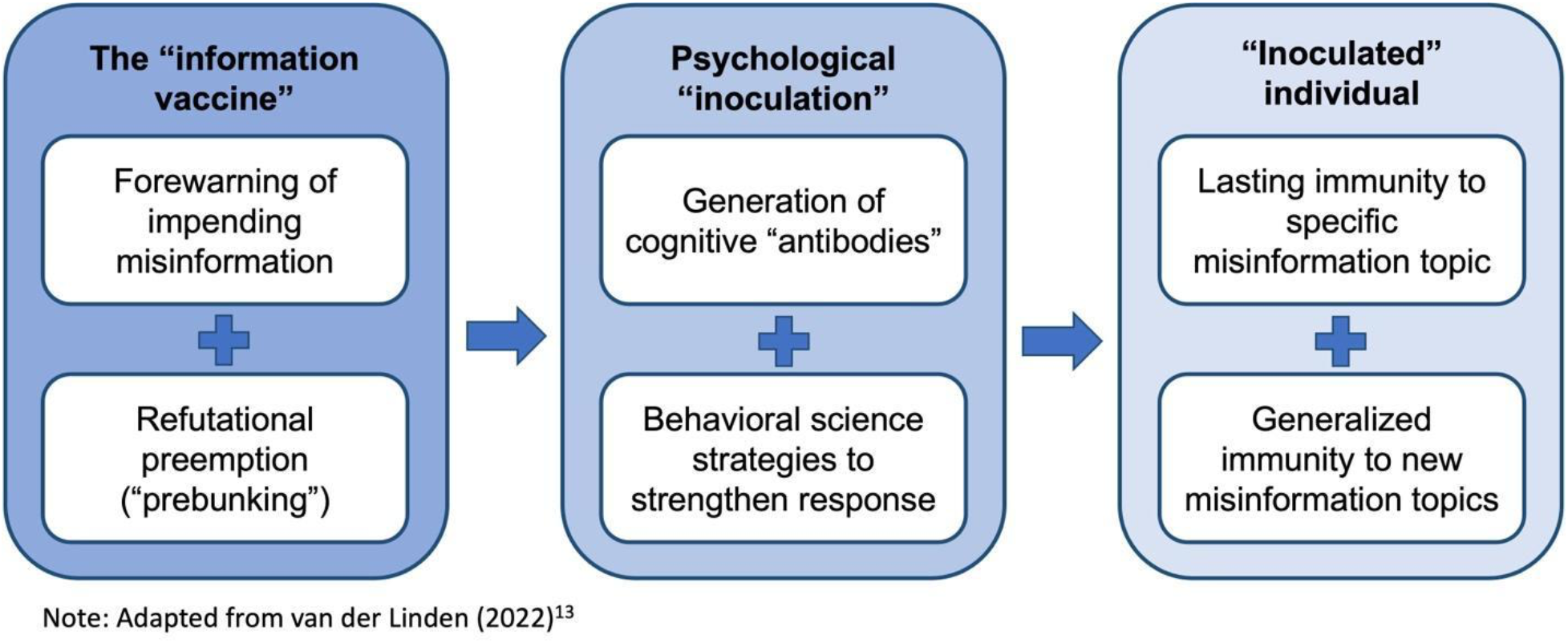
Psychological inoculation framework with behavioural science boost.

Prior research in other domains has shown that brief (5-15 minute) interactive prebunking interventions, often delivered via online games, can reduce belief in misinformation and reduce willingness to share them.^12^ Meta-analyses also support the efficacy of psychological inoculation.^12,13^ However, psychological inoculation has been studied primarily in high-income countries and for topics including COVID-19,^14,15^ climate change,^16^ and politics.^17^

There is limited evidence on the use of psychological inoculation for HIV-related misinformation, from low- and middle-income countries, and among young people.^18,19^ Furthermore, little is known about the effectiveness of brief, social-media-native video formats (e.g., TikTok’s distinct portrait-orientation reels) in delivering psychological inoculation at scale, which is an important gap in light of high social media penetration globally. Critical evidence gaps also persist on the durability of these effects,^10,20,21^ and the ability of inoculation interventions to confer generalised resistance to other forms of misinformation.

To address these gaps, we developed and tested two 2.5-minute videos that sought to inoculate viewers against common misinformation claims about a future HIV vaccine. We then conducted a pre-registered, two-arm randomised controlled trial with 2,163 women (18-29 years old) in South Africa. Our trial tested several pre-registered hypotheses: 1) participants who viewed the inoculation videos prior to exposure to misleading claims would report higher intentions to receive the HIV vaccine and be more likely to recommend the vaccine to a friend – an indicator of broader normative acceptance; 2) the inoculation videos would reduce the perceived credibility, emotional impact, and shareability of misinformation claims; 3) psychological inoculation would generalise to different misinformation claims and tactics not explicitly addressed in the videos; 4) the intervention would remain effective after 3-4 weeks.

## Methods

### Study design

We conducted a two-arm randomised controlled trial consisting of a main online survey (at study enrolment) and a follow-up survey approximately three weeks later. The main survey included: delivery of the intervention videos (or attention-matched control videos); exposure to multiple misinformation claims; pre- and post-intervention measurement of the primary outcome of vaccine intentions; and a study debrief highlighting that all misinformation claims were fabricated for research purposes. The follow-up survey included: exposure to additional misinformation claims; a re-assessment of vaccine intentions to examine the persistence of effects over time; and another study debrief. An outline of the study procedures and data collection is provided in appendix 1 (p 13, Figure S1). The study was reviewed and approved by independent research ethics committees at the University of Pennsylvania (854420), University of the Witwatersrand (reference number: 241118), University of Cape Town (reference number: 1018/2024), and Boston University (IRB number: H-45460).

### Study setting and procedures

We recruited participants from Prolific, an online research platform that includes thousands of individuals from South Africa.^22^ We enrolled participants who were female, aged 18-29 years, resident in South Africa, and able to read English (as indicated by Prolific as ‘fluent in English’); and had had sex in the past 12 months and reported their HIV status as negative or unknown. Although Prolific participants tend to be more educated than the general population in South Africa, the platform enabled rapid recruitment of a large, relevant cohort of young women who would be eligible for an HIV vaccine. Prolific invited eligible users to participate and provided a link to the study survey, which was administered through Qualtrics. Participants could complete the survey on any internet-enabled device.

After providing informed consent, participants answered questions related to socio-economic characteristics, mental health, and prior health behaviours. The survey also assessed participants’ HIV vaccine intentions and likelihood of recommending the HIV vaccine. To reduce the potential for demand-side characteristics to explain the results, all participants were provided with information about the HIV vaccine prior to randomisation to the intervention or control group: that any HIV vaccine available to the public in the future would have been tested and found to be safe and effective. Participants were then randomly assigned (1:1) to the intervention and control groups. Randomisation and survey administration were fully automated.

### Intervention group

Participants in the intervention group viewed two short inoculation videos, which were developed in three steps. First, we conducted a rapid review of misinformation claims about HIV vaccines (details published elsewhere),^23^ identifying concerns around side effects, fertility, and conspiracy theories. Second, we selected 54 unique claims from the review and surveyed young South African women (18–29 years; n=188) to determine which claims were most concerning to them. Safety-related claims — particularly those suggesting severe harm — were rated most concerning (details published elsewhere).^24^ Third, we developed a new set of false HIV-vaccine safety claims incorporating commonly used persuasion tactics when promoting misinformation.^25^ The claims mimic real-world statements that people could encounter. We tested their perceived credibility and influence on vaccine intentions and recommendations in a second online survey (n=195).

The two claims selected for the intervention scored highly in this second survey for how concerning, credible, and shareable they were; making them good candidates for inoculation. The first claim is “My sister got the HIV vaccine and her heart stopped the next day. Please don’t get vaccinated-IT COULD KILL YOU! 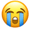”. This claim uses the post hoc fallacy, a misinformation tactic that exploits the human tendency to infer a cause-and-effect relationship based solely on the temporal order of events.^25^ The second claim is “HIV vaccine?! Yea, right, as soon as you can prove it’s ZERO risk! #HIVvaccine #nomorejabs”. This claim uses a misinformation tactic called “impossible expectations”, which is based on demanding unrealistic standards before acting on scientific evidence.^25^

Scripts for videos that inoculated against these two claims were developed following the three-part structure (see Figure 1): threat activation (alerting viewers that misinformation is common and about a specific false claim); refutation of the claim by explaining the common fallacies and manipulative tactics used in the claim; and a call to action (encouragement to look out for misinformation). The scripts were developed via participatory prototyping with young women (19-25 years old) in South Africa working as peer ambassadors for a mobile health clinic. Based on formative work assessing multiple modes or formats of inoculation content delivery (whiteboard animation, digital avatar, live video, or text), we then scripted and produced brief multi-character TikTok style videos filmed by a South African production company. Co-design workshops with peer health navigators and counselors from an HIV service delivery organisation helped ensure that the content, format, and actor choice were locally relevant, salient, and relatable. Inoculation Video 1 - post hoc fallacy – can be viewed at https://youtube.com/shorts/9Hs_36HS6Io?feature=share. Inoculation Video 2 - impossible expectations – can be viewed at https://youtube.com/shorts/viZUbEZJqfY?feature=share.

We also incorporated several strategies from the behavioural economics toolkit to strengthen message engagement, comprehension, retention, and to motivate action. Each video was followed by a short comprehension quiz (see appendix 2 (pp 16-19); incorrect answers required viewing the videos again since we sought to ensure that participants understood the tactics used in misinformation claims and thereby test the efficacy of psychological inoculation.^26^ Other strategies included cognitive engagement with the material through selection of a digital sticker at the end of the main survey; priming a new identity for the participants as misinformation spotters; and reminder SMS messages between the main and follow-up surveys to prompt sharing their new expertise in countering misinformation.

### Control group

Participants in the control arm viewed two attention-matched videos of identical length and style, and produced with the same actor, that focused on nutrition and diabetes, but with no inoculation content.

### Misinformation exposure and outcome measurement

After viewing the videos, all participants were shown the two primary misinformation claims – those directly addressed in the inoculation videos – presented as Facebook posts (see appendix 3 (p 20). Immediately after each post, participants rated (a) its credibility, (b) the extent to which it made them anxious or worried about an HIV vaccine, and (c) their likelihood of sharing it on social media. HIV vaccine intentions were then measured a second time to assess the impact of misinformation exposure on the primary outcome.

Next, participants viewed four additional misinformation claims and one true claim, presented in random order. For each claim, the same three outcomes – credibility, anxiety, and shareability – were assessed. The additional false claims were included to assess the extent to which the psychological inoculation might generalise beyond the HIV claims that were specifically inoculated against to new claims that used different content and tactics. HIV vaccine recommendation was measured again at the end of this sequence to capture changes after exposure to the full set of claims.

The follow-up survey was administered approximately three weeks later (median: 21 days; average: 22.6; SD: 5.4). Participants were shown the two primary misinformation claims again, along with four new misinformation claims and one new true claim, and completed the same set of outcome measures, including HIV vaccine intentions and HIV vaccine recommendation. The study ended with a second debrief identical to the main survey debrief.

### Outcomes

The primary outcome was participants’ intention to get vaccinated. Specifically, participants were asked: “If an HIV vaccine became available, how likely would you be to get vaccinated?” (1-10 scale, where 1 is extremely unlikely and 10 is extremely likely). Given that an HIV vaccine is not yet available, our measure proxied actual demand for the vaccine.

A secondary outcome assessed participants’ likelihood of recommending the vaccine to friends (“If an HIV vaccine became available, how likely would you be to recommend it to your friends?”; 1-10 scale). Additional secondary outcomes measured on 1-7 scales assessed the extent to which participants believed the claims (“How much do you believe this post is true?”); the anxiety induced by these claims (“How much does this post make you anxious or worried about the HIV vaccine?”); and the likelihood of sharing the claims (“If you saw this post on social media, how likely would you be to share it?”).

### Statistical analyses

We used ordinary least squares regression (OLS) models to estimate the effect of the intervention on primary and secondary outcomes, controlling for demographic, socioeconomic and other factors that have previously been shown to influence vaccine intentions and the perceived credibility of misinformation. These were participants’ age, race, education, province of residence (i.e., Gauteng or other), residential setting (i.e., informal settlement/township or other), employment status, and food insecurity. Province is relevant as the young people who co-designed the intervention resided in Gauteng province. While the intervention was designed for all young people in South Africa, it is possible it resonated more with people living in Gauteng. We also controlled for participant characteristics such as daily use of social media; symptoms of depression (i.e., PHQ-2 score ≥3); no receipt of a COVID-19 vaccine (proxy for being vaccine hesitant); ever used PrEP; HIV risk perception (binary variable identifying self-perceived high HIV risk); information avoidance behaviour (above the median); and self-reported confidence in scientists (1-4 scale). Finally, we also controlled for the pre-intervention values of the outcome variables in the regression specifications.

We also conducted pre-specified subgroup analyses to examine whether the effect of the intervention on study outcomes differed by age (<25 years vs. ≥25 years), prior PrEP use (never used vs. ever used), perceived risk of HIV (low vs. high), COVID-19 vaccination status (never received vs. received at least one dose), and information avoidance behaviour (low vs. high). For each sub-group characteristic, a separate analysis was conducted by including an interaction term between the treatment indicator and the corresponding binary subgroup indicator in a regression model that included the full sample. This interaction term tested whether the intervention effect differed significantly between a given subgroup and its complement. Effectiveness within a subgroup was defined as the magnitude and direction of the intervention effect in that subgroup, and a subgroup was characterised as showing greater effectiveness than its complement when the interaction term was statistically significant and indicated a larger treatment effect in that subgroup. All analyses were conducted using Stata version 16.1.

## Results

Between 24 July and 16 October 2025 we enrolled 2,163 participants, representing 93% of eligible individuals invited to participate (See CONSORT diagram, Figure 2). Participant characteristics were similar in the study groups (Table 1). Overall, the mean age was 25 years (standard deviation (SD): 2.57) and participants were predominantly Black African (91%). Education levels were high with 99% having completed high school (grade 12), but other measures indicated variation in socioeconomic status: 11% reported household hunger (past 7 days), and over a third were living in a township or informal settlement. Nearly all participants reported daily social media use and about half resided in Gauteng Province, the largest province in South Africa. Participants’ intentions to take the HIV vaccine were similar in both groups: mean of 7.81 and 7.77 in the control and intervention groups respectively.

**Figure 2.**
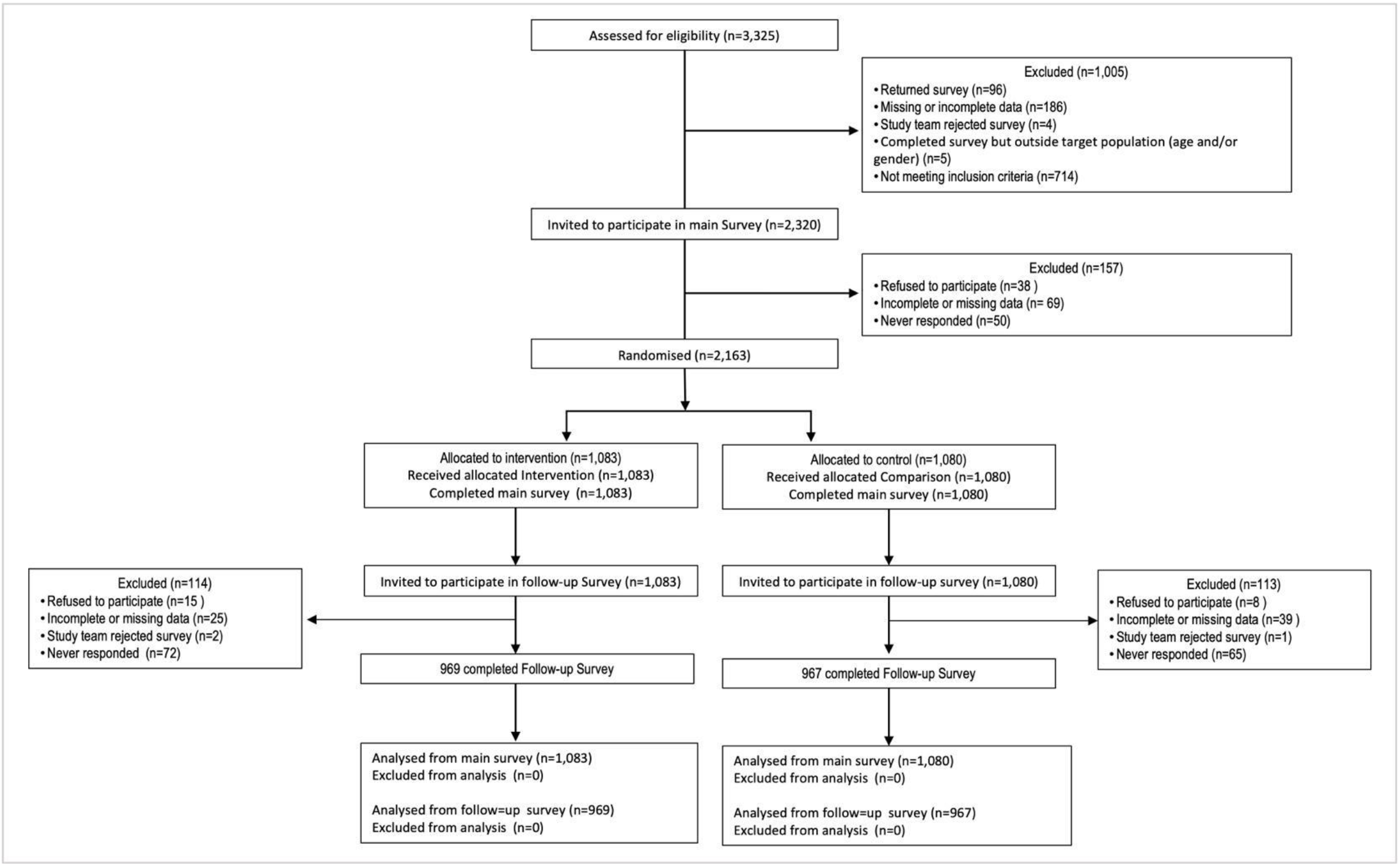
Consort diagram.

**Table 1.** Participant characteristics.

|  | All Participants | Control group<br>n (%) or<br>^Mean (SD) | Intervention group<br>n (%) or<br>^Mean (SD) |
|---|---|---|---|
| Demographics |  |  |  |
| Age | 25.1 (2.6)^ | 25.1 (2.6)^ | 25.1 (2.5)^ |
| African/Black | 1984 (91.7%) | 987 (91.4%) | 997 (92.1%) |
| Completed 12<sup>th</sup> grade in school | 2151 (99.4%) | 1071 (99.2%) | 1080 (99.7%) |
| Informal settlement/Township | 767 (35.5%) | 409 (37.9%) | 358 (33.1%) |
| Gauteng province resident | 1080 (49.9%) | 538 (49.8%) | 542 (50.0%) |
| Any wage work in last 7 days | 1790 (82.8%) | 890 (82.4%) | 900 (83.1%) |
| Information, Health Behaviours, and Psychosocial Factors |  |  |  |
| Any hunger in household in last 7 days | 243 (11.2%) | 124 (11.5%) | 119 (11.0%) |
| Any depression | 824 (38.1%) | 431 (39.9%) | 393 (36.3%) |
| Daily use of social media | 2119 (98.0%) | 1064 (98.5%) | 1055 (97.4%) |
| Never received COVID-19 vaccine | 546 (25.2%) | 257 (23.8%) | 289 (26.7%) |
| Ever used PrEP | 650 (30.1%) | 329 (30.5%) | 321 (29.6%) |
| Perceived risk of HIV is high<sup>#</sup> | 786 (36.3%) | 388 (35.9%) | 398 (36.8%) |
| Confidence in scientists [1-4] | 3.2 (0.6)^ | 3.2 (0.6)^ | 3.2 (0.6)^ |
| Any information avoidance behaviour | 1180 (54.6%) | 595 (55.1%) | 585 (54.0%) |
| Pre-intervention: Likely to get HIV vaccine [1-10] | 7.8 (2.8)^ | 7.8 (2.9)^ | 7.8 (2.8)^ |
| Pre-intervention: Likely to recommend HIV vaccine [1-10] | 8.5 (2.3)^ | 8.5 (2.4)^ | 8.6 (2.2)^ |
| Observations | 2,163 | 1,080 | 1,083 |

Notes: Continuous variables are indicated with ^ and reported as means (standard deviation), while all other variables are binary and reported as n (%). SD: Standard deviation. <sup>#</sup>Perceived high (vs low) HIV risk is defined as responding “moderate” or “great” to the question: “Do you think you have no risk, a small risk, a moderate risk or a great risk of getting the HIV virus?”

HIV vaccine intentions decreased in the control group after seeing the misinformation claims (−0.63, SD:1.92). In contrast, intentions to take the vaccine increased in the intervention group after seeing the inoculation video and then being shown the misinformation claims (+0.30, SD:1.82). See Figure S2. Controlling for covariates and baseline vaccine intentions, vaccine intentions post misinformation exposure was 0.92 (12%) higher (p<0.01; 95%CI: 0.78,1.06) in the intervention group compared to the control group (Figure 3 Panel A and appendix 1 (p 1, Table S1).

**Figure 3.**
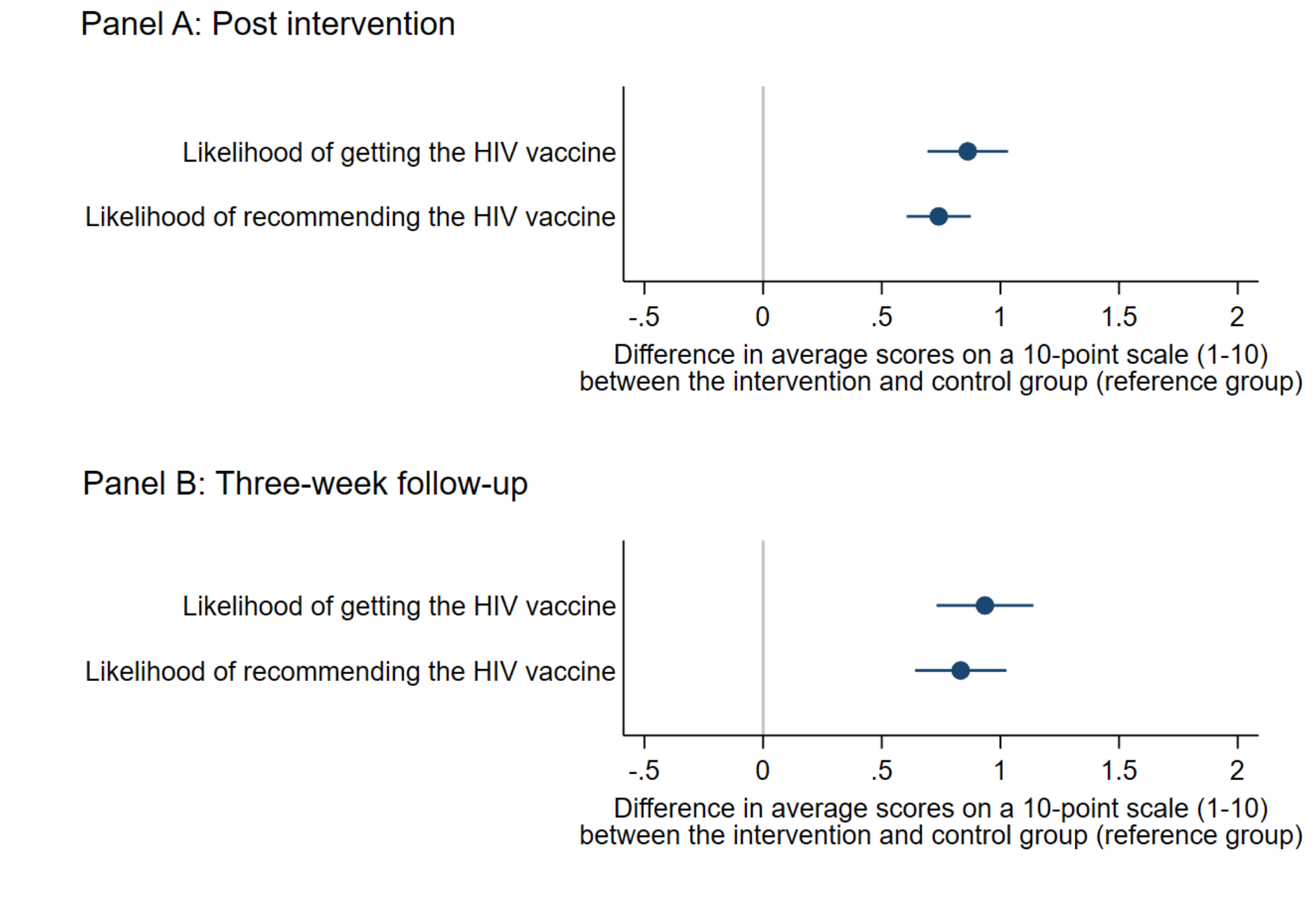
Effects of inoculation videos on vaccine intentions and vaccine promotion immediately post intervention and after three weeks. Post-intervention (Panel A) and three-weeks follow-up (Panel B) effects of the inoculation videos on HIV vaccine intentions and likelihood of recommending the HIV vaccine to a friend were estimated using linear regression models controlling for preintervention measures of the outcome and covariates (using data collected during the main survey at study enrolment). See appendix 1 (p 1-2, Tables S1 (vaccine intentions) and S2 (vaccine recommendation)) for all coefficients.

Similarly, participants’ self-reported likelihood of recommending the HIV vaccine to a friend declined in the control group after seeing the misinformation claims (−0.63, SD:1.90), whereas they increased in the intervention group (+0.11, SD:1.63). See Figure S3. Controlling for covariates (Figure 3 Panel A and appendix 1 (p 2, Table S2), intervention group participants were more likely to recommend the HIV vaccine to a friend than control group participants (+0.74, p<0.01, 95%CI: 0.61,0.88).

Sensitivity analysis excluding individuals who answered quiz questions incorrectly and were asked to rewatch either video (n=29), found substantively similar results (appendix 1 (pp 3-4, Table S3 and S4).

Inoculation messages also reduced (32-55%) how much participants believed all six false claims shown in the main survey (Figure 4 Panel A, appendix 1 (p 5, Table S5): someone’s heart stopped after getting the HIV vaccine (Claim #1, specifically inoculated against, −1.7, p<0.01); that you should not get the HIV vaccine unless it is 100% safe (Claim #2, specifically inoculated against, −2.2, p<0.01); the HIV vaccine causes someone to get HIV (Claim #3, −1.0, p<0.01); the HIV vaccine is not worth getting unless 100% effective (Claim #4, −2.0, p<0.01); the COVID-19 vaccine was created by the government to sterilise Black women (Claim #5, −0.6, p<0.01); and a person should not get any vaccine unless there are no side-effects (Claim #6, −2.4, p<0.01).

**Figure 4.**
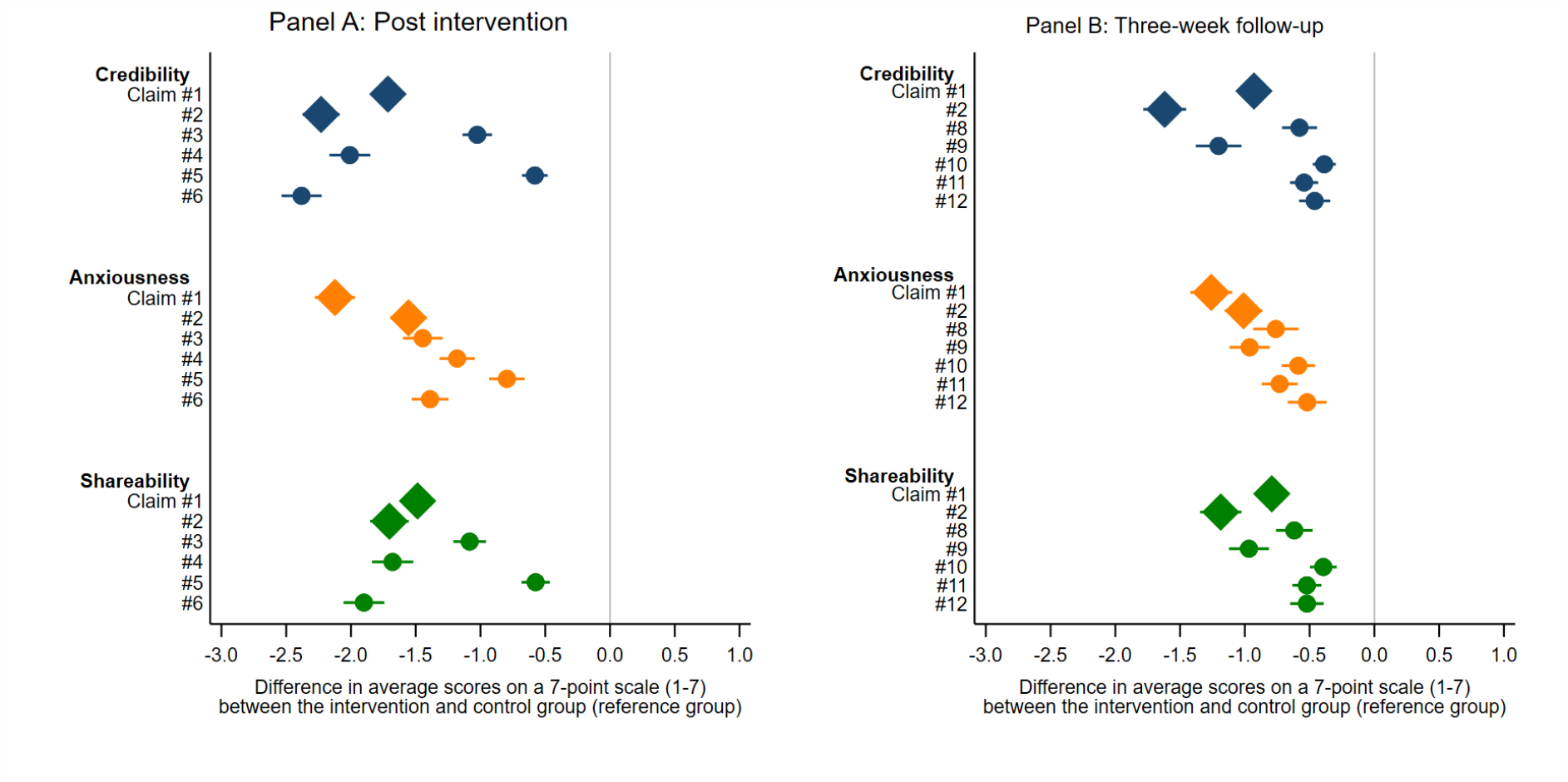
Effects of the inoculation videos on credibility, emotional response to, and shareability of misinformation claims immediately post intervention and after three weeks. The effects of the inoculation videos on the perceptions of each claim’s credibility, the extent to which the claim made the participant anxious/nervous, and the likelihood that they would share the claim if they saw it on social media were estimated using linear regression models controlling for covariates and using data from the main survey at study enrolment. Coefficients for the two primary misinformation claims (claim #1 and claim #2, i.e. those directly addressed in the inoculation videos) are represented with diamonds. <u>Panel A</u> - post intervention, main survey: Summary of each claim: Claim #1: someone’s heart stopped after getting the HIV vaccine;”’ “Claim #2: you should not get the HIV vaccine unless it is 100% safe; Claim #3: the HIV vaccine causes someone to get HIV; Claim” “#4: the HIV vaccine is not worth getting unless 100% effective; Claim #5: the COVID-19 vaccine was created by the government to” “sterilise Black women; Claim #6: a person should not get any vaccine unless there are no side-effects.” See appendix 3 (p 20) for the exact wording for each claim. See appendix 1 (p 5) See appendix 1 (p 5) Table S5 for all post-intervention model coefficients. <u>Panel B</u> - follow-up survey: Summary of each claim: Claim #1: someone’s heart stopped after getting the HIV vaccine;”’ “Claim #2: you should not get the HIV vaccine unless it is 100% safe; Claim #3: the HIV vaccine causes someone to get HIV; Claim” “#4: the HIV vaccine is not worth getting unless 100% effective; Claim #5: the COVID-19 vaccine was created by the government to” “sterilise Black women; Claim #6: a person should not get any vaccine unless there are no side-effects.” See appendix 3 (p 20) for the exact wording for each claim. See appendix 1 (p 7) Table S7 for all three-week follow-up model coefficients.

The inoculation videos reduced negative emotional reactions to, and reduced likelihood of sharing the misinformation (a behavioural indicator of intent to engage with and propagate misinformation).^13^ Compared to the control group, the intervention group reported feeling less anxious or worried about the claim (33-49% lower), with the difference in average scores ranging from −0.80 (p<0.01, 95% CI: −0.93, −0.66) to −2.12 (p<0.01, 95% CI: −2.28, −1.97). Intervention group reported being less likely than the control group to share the six misinformation claims with others if they saw them on social media (range: *β*:-0.57 (p<0.01, 95% CI: −0.68, −0.46) to *β*:-1.90 (p<0.01, 95% CI: −2.06, −1.74)).

The 3-week follow-up survey re-interviewed 1,936 (89.5%) participants, with attrition similar in the intervention group (114/1083 (10.5%)) and control group (113/1080 (10.5%)). At 3-week follow-up, the intervention group continued to report stronger intentions to accept an HIV vaccine than the control group after viewing additional misinformation claims (+0.97, p<0.01, 95% CI: 0.78, 1.17). Descriptive analysis showed that in the control group, HIV vaccine intentions were reduced by 13% (from 7.8/10 to 6.8/10), while in the treatment group there was no change in vaccine intentions (7.8/10 vs 7.8/10). The intervention group remained more likely to recommend the HIV vaccine to a friend than the control group (*β*: 0.83, p<0.01, 95% CI: 0.64, 1.03) (Figure 3 Panel B); continued to perceive misinformation to be less credible, were less likely to report that misinformation made them anxious or worried, and were less likely to share misinformation (Figure 4 Panel B). A logistic regression analysis of factors associated with attrition between the surveys found that no covariates were significantly associated with attrition (appendix 1 (p 6, Table S6).

To assess the possibility that the inoculation intervention also increased skepticism about true claims,^12^ we asked participants to view a true claim in the main survey (that the vaccine will only be offered to the public once it is tested and approved) and evaluate its credibility. The intervention group expressed slightly higher (rather than lower) belief in the true statement than the control group (5.59 vs 5.75; *β*: 0.16, p<0.05, 95% CI: 0.02, 0.30). The follow-up survey also assessed responses to a true claim: “Smoking literally increases your risk for lung cancer for sure −- that’s been proven! But that doesn’t mean everyone who smokes gets lung cancer.” Counter to expectations, the intervention group was less likely to believe this claim than the control group (*β*: −0.29, p<0.01, 95% CI: −0.47,-0.11).

Pre-specified sub-group analyses (see appendix 1 (pp 8-11, Tables S8-S11) found that the inoculation messages were most effective among participants who had not received the COVID-19 vaccine at enrolment: HIV vaccine intentions in the follow-up survey (*β* for interaction term: +0.83, p<0.01, 95% CI: 0.39, 1.30); HIV vaccine recommendation at main survey (*β* for interaction term: +0.56, p<0.01, 95% CI: 0.25, 0.87); and HIV vaccine recommendation at follow-up (*β* for interaction term: +0.75, p<0.01, 95% CI: 0.31, 1.19). For three other pre-specified sub-groups we found little evidence of subgroup-specific intervention effects: age, PrEP use, perceived HIV risk, and information avoidance. An additional subgroup analysis conducted to assess potential differences in intervention effect by socioeconomic status found that our intervention had similar effects on HIV-vaccine intentions among those residing in an informal settlement/township and those living elsewhere (see appendix 1 (p 12, Table S12).

## Discussion

In this randomised controlled trial with young women in South Africa, brief psychological inoculation messages delivered through short videos were effective in protecting participants from the negative effects of misinformation about a yet-to-be-developed HIV vaccine. The inoculation messages prevented the substantial decline in intentions to accept an HIV vaccine observed in the control group after encountering misinformation. The intervention also reduced the perceived credibility of multiple misinformation claims, lowered negative emotional reactions to misinformation, and decreased participants’ likelihood of sharing false claims. Importantly, these protective effects persisted three weeks later, even after participants saw a broader range of misinformation claims. Together, these findings show that inoculation messages can meaningfully preserve vaccine intentions and reduce susceptibility to misinformation in a population particularly at risk of acquiring HIV.

Furthermore, the protective effects of the inoculation videos extended beyond the two specific claims about the HIV vaccine addressed in the videos. The additional claims included ones using the same persuasion tactics as in the main claims (post hoc fallacy and impossible expectations) but with different content and in relation to vaccines in general, the flu vaccine, and different health topics. They also included claims using different persuasion tactics and about other vaccines or HIV prevention methods. Inoculated participants also rated these new claims as less credible, experienced lower levels of worry in response to them, and reported a lower likelihood of sharing them compared to the control group. This is an encouraging finding given how rapidly false claims evolve and how difficult it is to pre-emptively counter every claim that may emerge. It points to the potential for concise, scalable inoculation interventions to offer broader protection against misinformation.

While we found no evidence that the inoculation videos made participants more suspicious of accurate health claims immediately after receiving the intervention, at the 3-week follow-up the intervention group was slightly less likely than the control group to believe that smoking increases risk of lung cancer but that not everyone who smokes gets lung cancer. This result could indicate a small backfire effect of inoculation that should be considered in future studies and intervention delivery. One explanation for this finding, however, is that the claim included language that might have been seen to be consistent with the persuasion technique of impossible expectations (“increases your risk of lung cancer for sure”), which was the focus of the second inoculation video. The intervention group may have become more skeptical about posts that make absolute claims. Another explanation is that the inoculation videos’ call to action – to trust only credible sources – may have led participants to distrust a Facebook post by “Slay Queen.” Finally, there is potential that the intervention group found this claim suspicious given the true claim had nothing to do with the intervention or misinformation topic (the HIV vaccine). Our main survey results are consistent with findings of a recent meta-analysis,^12^ which found that psychological inoculation improved the ability to identify manipulative content specifically rather than increasing generalized scepticism.

There are several mechanisms through which brief, engaging inoculation videos may have combatted misinformation. First, consistent with the inoculation (prebunking) approach and given the HIV vaccine is not available yet, participants most likely had not yet formed strong opinions or preferences related to the HIV vaccine nor had been exposed to many claims about it. This highlights the importance of getting out ahead of misinformation when a new health technology is being introduced. Second, the protection/immunity found in our study beyond the specific claims we inoculated against is consistent with the theoretical proposition that inoculation against underlying argumentative tactics, rather than against specific factual assertions, can build a broader cognitive immune response.^12^ Third, the intervention videos used a familiar, highly engaging social-media format and incorporated multiple behavioural insights to increase attention, salience, and relatability. This points to the importance of ensuring the potency and relatability of brief video interventions like ours to enable broad dissemination.

To our knowledge, this is the first psychological inoculation study to focus specifically on HIV prevention in sub-Saharan Africa – an important context given the high burden of HIV and the rapid diffusion of health misinformation through social media. Our findings contribute to an emerging literature on the effectiveness of prebunking interventions in mitigating the influence of misinformation, while extending this literature into the domain of HIV prevention in a middle-income country setting. Two previous studies have assessed the impact of psychological inoculation on condom use and disclosure among people living with HIV.^18,19^ While other inoculation studies demonstrating significant effects typically have more intensive and active inoculation (e.g., playing an interactive game), our light touch, passive inoculation intervention (watching two 2.5-minute videos) achieved similar results in a format that is easily scalable. Our results align with another study evaluating short inoculation videos: in standardized terms, our effect (Cohen’s d = 0.33) falls within the range reported across the seven inoculation video substudies targeting misinformation more broadly (d = 0.28–0.68).^27^ The durability of the intervention’s effects over a three-week period also compare favourably to other inoculation studies,^10,12,20^ and could potentially be considered an underestimation if the study debrief at the end of the main survey increased scepticism of misinformation among both the control group and treatment group.

This study has several limitations. First, we assessed participants’ vaccine intentions rather than actual health behaviour. Because an HIV vaccine does not yet exist, measuring stated demand is the most feasible proxy for potential uptake, and the hypothetical context is valid for the intervention and control groups. The use of intention measures is common in online experimental studies, but the extent to which intentions translate into real-world behaviour remains uncertain. We do however know that the study intervention reduced how much people believed misinformation, and misinformation has been found to be associated with reduced vaccine uptake.^5,28^ Second, participants were recruited through an online research platform and may not fully reflect the broader population of adolescent girls and young women in South Africa. Although nearly all participants were daily social media users the generalisability of our findings to offline or harder-to-reach populations requires further study. Our findings may nonetheless apply more broadly as psychological inoculation effects have been found to be similar across education levels.^29^ In addition, we found that our intervention had similar effects on HIV-vaccine intentions among those residing in an informal settlement/township and those living elsewhere. This finding provides some confidence that results may apply to young women across a broader socioeconomic spectrum. We also found that 36.3% of our study participants reported a “moderate” or “great risk” of HIV infection, indicating our sample is the population of interest when rolling out HIV prevention services. Third, we cannot isolate the effect of the inoculation videos completely from the effect of the other intervention components. One concern with the main study findings is that this could be driven by intervention participants answering the quiz questions incorrectly and being asked to rewatch the video. We are confident that the effects found in the main survey are caused by the intervention given that only 29 participants were asked to rewatch the video, and that the main survey results are robust to the exclusion of these participants. The identity prime at the end of the main survey and SMS reminder before the follow-up might have boosted the intervention effect slightly at follow-up. While future research should isolate the inoculation impact, it is likely that nudges such as these have relatively small effects in comparison to the main intervention. Fourth, our follow-up period was limited to approximately three weeks (which is longer than many inoculation studies that do not include a follow-up). While we observed encouraging durability over this interval, longer-term assessments are needed to determine whether the protective effects of inoculation persist as misinformation evolves. Fifth, while the control group received the same information about the HIV vaccine as provided to the intervention group in the inoculation videos – any HIV vaccine available to the public in the future would have been tested and found to be safe and effective – it is possible that the results could, in part, be explained by demand-side characteristics. Sixth, we do not know whether our results would generalise to other country contexts with lower prevalence of HIV.

Although an HIV vaccine is not yet available, there are ongoing efforts to develop a safe and effective vaccine. Equipping people with the ability to detect and reject false claims before health products become widely available is essential for ensuring high demand, given mounting evidence that debunking misinformation after it has spread is much more difficult and often less effective.^30^ Our findings are also extremely timely in light of the imminent rollout of lenacapavir, a groundbreaking long-acting medication for HIV prevention that is very likely to be the subject of considerable misinformation about its safety, biological mechanisms, and the populations for which it is intended. Investing early in the participatory design, adaptation, and dissemination of scalable prebunking interventions may help protect public confidence and boost demand for lenacapavir and other novel HIV prevention technologies.

## Supporting information

Appendix Tables, Figures, Quiz and Misinformation Claims

## Data Availability

All data produced in the present study are available upon reasonable request to the authors.

## Acknowledgements

This research was supported by the National Institutes of Health [1R01MH132401-01]. The views and content are solely the responsibility of the authors and do not represent the official views, policies or guidelines of any funder. The intervention videos were produced by Reel Epics Productions, an award-winning production company based in South Africa. Peer Ambassadors from Shout-It-Now co-designed the intervention videos. Megan Rabin provided valuable input into the selection of the misinformation claims and study methods.

## Data availability

The data that support the findings of this study will be made available in de-identified form upon request to the authors. A detailed data dictionary and all materials necessary to reproduce the analyses will also be provided.

## Contributions

Conceptualisation: A.B., H.T., B.M-B., L.L. C.C-M., S.P.; data curation: B.M-B., A.B., D.P.; formal analysis: D.P., B.M-B., H.T., A.B., A.S-S.; funding acquisition: A.B., H.T., B.M-B.; investigation: B.M-B., D.P., S.B., T.E., L-G.B., C.C-M., L.L., S.P., A.S-S., S.v.d.L, H.T., A.B.; methodology: B.M-B., S.B., T.E., C.C-M., L.L., S.P., A.S-S., S.v.d.L, H.T., A.B.; project administration: A.B., H.T., B.M-B.; supervision: A.B., H.T., B.M-B.; visualisation: A.B., D.P., B.M-B., H.T.; writing—original draft: B.M-B., D.P., H.T., A.B.; writing—review and editing: B.M-B., D.P., S.B., T.E., L-G.B., C.C-M., L.L., S.P., A.S-S., S.v.d.L, H.T., A.B. A.B., H.T., B.M-B. had full access to all the data in the study and had final responsibility for the decision to submit for publication.

## Ethics declarations

The study was reviewed and approved by independent research ethics committees at the University of Pennsylvania (854420), University of the Witwatersrand (reference number: 241118), University of Cape Town (reference number: 1018/2024), and Boston University (IRB number: H-45460). The study procedures were developed with guidance and monitoring from a four-person data safety and monitoring board with expertise in randomised controlled trials, epidemiology and vaccine research. The study was also preregistered on ClinicalTrials.Gov (NCT06700447) and the South African National Clinical Trials Register (DOH-27-032025-5053).

