## Appendix Tables, Figures, Quiz and Misinformation Claims for "Psychological inoculation protects young women in South Africa against HIV prevention misinformation: Evidence from a randomised controlled trial"

### Supplementary Appendix 1: Tables and Figures

**Article**: Psychological inoculation protects young women in South Africa against HIV prevention misinformation: Evidence from a randomised controlled trial

**Table S1. OLS Estimates of the Intervention Effect on Post-Inoculation Vaccine Intentions Among Women Aged 18–29 in South Africa**

| | Main survey | | | | Follow-up survey | | | |
|---|---|---|---|---|---|---|---|---|
| | (1) | (2) | (3) | (4) | (5) | (6) | (7) | (8) |
| Intervention | 0.893*** (0.667 - 1.120) | 0.920*** (0.776 - 1.064) | 0.921*** (0.777 - 1.065) | 0.917*** (0.776 - 1.058) | 1.006*** (0.755 - 1.256) | 0.990*** (0.787 - 1.193) | 0.981*** (0.778 - 1.184) | 0.970*** (0.775 - 1.165) |
| Pre-inoculation vaccine intention | | 0.731*** (0.706 - 0.756) | 0.727*** (0.702 - 0.753) | 0.664*** (0.636 - 0.692) | | 0.577*** (0.541 - 0.612) | 0.569*** (0.532 - 0.605) | 0.463*** (0.424 - 0.502) |
| Demographic covariates | No | No | Yes | Yes | No | No | Yes | Yes |
| Information and health behaviours covariates | No | No | No | Yes | No | No | No | Yes |
| Observations | 2,163 | 2,163 | 2,163 | 2,163 | 1,936 | 1,936 | 1936 | 1,936 |
| R-squared | 0.027 | 0.609 | 0.610 | 0.629 | 0.031 | 0.365 | 0.368 | 0.423 |
| Control-group mean | 7.178 | 7.178 | 7.178 | 7.178 | 6.818 | 6.818 | 6.818 | 6.818 |

95% CI in parentheses
*** p<0.01, ** p<0.05
Outcome: Post-inoculation HIV vaccine intentions measured on a scale between 1-10, where 1 corresponds to the lowest intention of getting vaccinated and 10 the highest.
Demographic covariates include age, African/Black race, completion of 12th grade, residence in an informal settlement or township, Gauteng province residence, and any wage work in the past seven days. Information and health behaviour covariates include any household hunger in the past seven days, any depression, daily social media use, never having received a COVID-19 vaccine, ever having used PrEP, high perceived HIV risk, confidence in scientists (1–4 scale), and any information avoidance behaviour.

**Table S2. OLS Estimates of the Intervention Effect on Post-Inoculation Vaccine Recommendations Among Women Aged 18–29 in South Africa**

| | Main Survey | | | | Follow-up Survey | | | |
|---|---|---|---|---|---|---|---|---|
| | (1) | (2) | (3) | (4) | (5) | (6) | (7) | (8) |
| Intervention | 0.772*** | 0.752*** | 0.750*** | 0.740*** | 0.906*** | 0.852*** | 0.844*** | 0.833*** |
| | (0.576 - 0.968) | (0.613 - 0.891) | (0.611 - 0.889) | (0.605 - 0.875) | (0.668 - 1.143) | (0.650 - 1.053) | (0.643 - 1.045) | (0.640 - 1.026) |
| Pre-inoculation vaccine recommendation | | 0.714*** | 0.707*** | 0.637*** | | 0.610*** | 0.597*** | 0.481*** |
| | | (0.684 - 0.744) | (0.676 - 0.737) | (0.604 - 0.670) | | (0.566 - 0.653) | (0.553 - 0.641) | (0.435 - 0.528) |
| Demographic covariates | No | No | Yes | Yes | No | No | Yes | Yes |
| Information and health behaviours covariates | No | No | No | Yes | No | No | No | Yes |
| Observations | 2,163 | 2,163 | 2,163 | 2,163 | 1,936 | 1,936 | 1,936 | 1,936 |
| R-squared | 0.027 | 0.511 | 0.515 | 0.543 | 0.028 | 0.301 | 0.307 | 0.367 |
| Control-group mean | 7.892 | 7.892 | 7.892 | 7.892 | 7.212 | 7.212 | 7.212 | 7.212 |

95% CI in parentheses
*** p<0.01, ** p<0.05
Outcome: Post-inoculation vaccine recommendation measured on a scale between 1-10, where 1 corresponds to the lowest likelihood of recommending the vaccine to friends and 10 the highest.
Demographic covariates include age, African/Black race, completion of 12th grade, residence in an informal settlement or township, Gauteng province residence, and any wage work in the past seven days. Information and health behaviour covariates include any household hunger in the past seven days, any depression, daily social media use, never having received a COVID-19 vaccine, ever having used PrEP, high perceived HIV risk, confidence in scientists (1–4 scale), and any information avoidance behaviour.

**Table S3. OLS Estimates of the Intervention Effect on Post-Inoculation Vaccine Intentions Among Women Aged 18–29 in South Africa, excluding participants (n=29) who watched either intervention video twice**

| | Main survey | | | | Follow-up survey | | | |
|---|---|---|---|---|---|---|---|---|
| VARIABLES | (1) | (2) | (3) | (4) | (5) | (6) | (7) | (8) |
| Intervention | 0.910*** | 0.932*** | 0.932*** | 0.932*** | 1.004*** | 0.985*** | 0.985*** | 0.985*** |
| | (0.682 - 1.137) | (0.788 - 1.076) | (0.788 - 1.076) | (0.788 - 1.076) | (0.752 - 1.256) | (0.780 - 1.189) | (0.780 - 1.189) | (0.780 - 1.189) |
| Pre-inoculation vaccine intention | | 0.728*** | 0.728*** | 0.728*** | | 0.578*** | 0.578*** | 0.578*** |
| | | (0.703 - 0.753) | (0.703 - 0.753) | (0.703 - 0.753) | | (0.542 - 0.614) | (0.542 - 0.614) | (0.542 - 0.614) |
| Demographic covariates | No | No | Yes | Yes | No | No | Yes | Yes |
| Information and health behaviours covariates | No | No | No | Yes | No | No | No | Yes |
| Observations | 2,134 | 2,134 | 2,134 | 2,134 | 1,911 | 1,911 | 1,911 | 1,911 |
| R-squared | 0.028 | 0.609 | 0.609 | 0.609 | 0.031 | 0.365 | 0.365 | 0.365 |
| Control-group mean | 7.178 | 7.178 | 7.178 | 7.178 | 6.818 | 6.818 | 6.818 | 6.818 |

95% CI in parentheses
*** p<0.01, ** p<0.05
Outcome: Post-inoculation HIV vaccine intentions measured on a scale between 1-10, where 1 corresponds to the lowest intention of getting vaccinated and 10 the highest. Demographic covariates include age, African/Black race, completion of 12th grade, residence in an informal settlement or township, Gauteng province residence, and any wage work in the past seven days. Information and health behaviour covariates include any household hunger in the past seven days, any depression, daily social media use, never having received a COVID-19 vaccine, ever having used PrEP, high perceived HIV risk, confidence in scientists (1–4 scale), and any information avoidance behaviour.

**Table S4. OLS Estimates of the Intervention Effect on Post-Inoculation Vaccine Recommendations Among Women Aged 18–29 in South Africa, excluding participants (n=29) who watched either intervention video twice**

| | Main survey | | | | Follow-up survey | | | |
|---|---|---|---|---|---|---|---|---|
| VARIABLES | (1) | (2) | (3) | (4) | (5) | (6) | (7) | (8) |
| Intervention | 0.778*** | 0.762*** | 0.762*** | 0.762*** | 0.913*** | 0.862*** | 0.862*** | 0.862*** |
| | (0.581 - 0.975) | (0.623 - 0.901) | (0.623 - 0.901) | (0.623 - 0.901) | (0.675 - 1.151) | (0.660 - 1.064) | (0.660 - 1.064) | (0.660 - 1.064) |
| Pre-inoculation vaccine recommendation | | 0.714*** | 0.714*** | 0.714*** | | 0.607*** | 0.607*** | 0.607*** |
| | | (0.684 - 0.744) | (0.684 - 0.744) | (0.684 - 0.744) | | (0.563 - 0.650) | (0.563 - 0.650) | (0.563 - 0.650) |
| Demographic covariates | No | No | Yes | Yes | No | No | Yes | Yes |
| Information and health behaviours covariates | No | No | No | Yes | No | No | No | Yes |
| Observations | 2,134 | 2,134 | 2,134 | 2,134 | 1,911 | 1,911 | 1,911 | 1,911 |
| R-squared | 0.027 | 0.515 | 0.515 | 0.515 | 0.029 | 0.302 | 0.302 | 0.302 |
| Control-group mean | 7.892 | 7.892 | 7.892 | 7.892 | 7.212 | 7.212 | 7.212 | 7.212 |

95% CI in parentheses
*** p<0.01, ** p<0.05
Outcome: Post-inoculation vaccine recommendation measured on a scale between 1-10, where 1 corresponds to the lowest likelihood of recommending the vaccine to friends and 10 the highest. Demographic covariates include age, African/Black race, completion of 12th grade, residence in an informal settlement or township, Gauteng province residence, and any wage work in the past seven days. Information and health behaviour covariates include any household hunger in the past seven days, any depression, daily social media use, never having received a COVID-19 vaccine, ever having used PrEP, high perceived HIV risk, confidence in scientists (1–4 scale), and any information avoidance behaviour.

**Table S5. OLS Estimates of the Intervention Effect on Self-Reported Credibility, Anxiety, and Willingness to Share Misinformation Claims Among Women Aged 18–29 in South Africa in the Main Survey**

| | (1)<br>Claim 1 | (2)<br>Claim 2 | (3)<br>Claim 3 | (4)<br>Claim 4 | (5)<br>Claim 5 | (6)<br>Claim 6 | (7)<br>Claim 7^ |
|---|---|---|---|---|---|---|---|
| | | | | Credibility (1-7 scale) | | | |
| Intervention | -1.714***<br>(-1.825 - - 1.603) | -2.231***<br>(-2.374 - - 2.087) | -1.025***<br>(-1.139 - - 0.911) | -2.008***<br>(-2.167 - - 1.850) | -0.580***<br>(-0.680 - - 0.481) | -2.381***<br>(-2.536 - - 2.226) | 0.159**<br>(0.025 - 0.293) |
| Covariates | Yes | Yes | Yes | Yes | Yes | Yes | Yes |
| Observations | 2,163 | 2,163 | 2,163 | 2,163 | 2,163 | 2,163 | 2,163 |
| R-squared | 0.323 | 0.322 | 0.180 | 0.243 | 0.112 | 0.315 | 0.044 |
| Control-group mean | 3.096 | 4.197 | 2.351 | 4.156 | 1.840 | 4.418 | 5.590 |
| | | | | Anxiousness (1-7 scale) | | | |
| Intervention | -2.123***<br>(-2.279 - - 1.967) | -1.555***<br>(-1.696 - - 1.415) | -1.446***<br>(-1.599 - - 1.292) | -1.180***<br>(-1.317 - - 1.044) | -0.796***<br>(-0.933 - - 0.659) | -1.389***<br>(-1.532 - - 1.247) | -0.638***<br>(-0.784 - - 0.492) |
| Covariates | Yes | Yes | Yes | Yes | Yes | Yes | Yes |
| Observations | 2,163 | 2,163 | 2,163 | 2,163 | 2,163 | 2,163 | 2,163 |
| R-squared | 0.284 | 0.222 | 0.193 | 0.170 | 0.120 | 0.189 | 0.071 |
| Control-group mean | 4.315 | 3.407 | 3.186 | 3.014 | 2.443 | 3.319 | 2.516 |
| | | | | Shareability (1-7 scale) | | | |
| Intervention | -1.486***<br>(-1.619 - - 1.352) | -1.703***<br>(-1.852 - - 1.554) | -1.083***<br>(-1.209 - - 0.957) | -1.678***<br>(-1.838 - - 1.519) | -0.574***<br>(-0.684 - - 0.464) | -1.900***<br>(-2.057 - - 1.742) | |
| Covariates | Yes | Yes | Yes | Yes | Yes | Yes | |
| Observations | 2,163 | 2,163 | 2,163 | 2,163 | 2,163 | 2,163 | |
| R-squared | 0.223 | 0.228 | 0.161 | 0.192 | 0.105 | 0.243 | |
| Control-group mean | 2.889 | 3.353 | 2.402 | 3.453 | 1.893 | 3.622 | |

95% CI in parentheses
*** p<0.01, ** p<0.05
Outcomes: Credibility is measured on a scale of 1-7, where 1 corresponds to the lowest self-reported likelihood of believing the misinformation claim and 7 the highest. Anxiousness is measured on a scale of 1-7, where 1 corresponds to the lowest self-reported anxiety upon seeing the misinformation claim and 7 the highest. Shareability is measured on a scale of 1-7, where 1 corresponds to the lowest self-reported likelihood of sharing the misinformation claim and 7 the highest.
Demographic covariates include age, African/Black race, completion of 12th grade, residence in an informal settlement or township, Gauteng province residence, and any wage work in the past seven days. Information and health behaviour covariates include any household hunger in the past seven days, any depression, daily social media use, never having received a COVID-19 vaccine, ever having used PrEP, high perceived HIV risk, confidence in scientists (1–4 scale), and any information avoidance behaviour.
^Claim 7 is the ‘true’ claim that presented information considered legitimate.
Data on the shareability of Claim 7 was not collected due to a survey error.

**Table S6. Logistic Regression Estimates of Characteristics Associated with Attrition from the Follow-Up Survey**

| | |
|---|---|
| Age | 0.008<br>(-0.046 - 0.063) |
| African/Black | -0.442<br>(-0.915 - 0.031) |
| Completed 12th grade in school | 0.429<br>(-1.644 - 2.502) |
| Informal settlement/Township | 0.098<br>(-0.195 - 0.391) |
| Gauteng province resident | -0.114<br>(-0.396 - 0.168) |
| Any wage work in last 7 days | -0.000<br>(-0.374 - 0.374) |
| Any hunger in household in last 7 days | -0.452<br>(-1.285 - 0.381) |
| Any depression | 0.072<br>(-0.216 - 0.360) |
| Daily use of social media | 0.184<br>(-0.124 - 0.493) |
| Never received COVID-19 vaccine | 0.034<br>(-0.259 - 0.327) |
| Ever used PrEP | -0.182<br>(-0.431 - 0.066) |
| Perceived risk of HIV is high | -0.092<br>(-0.386 - 0.203) |
| Confidence in scientists [1-4] | 0.104<br>(-0.322 - 0.530) |
| Any information avoidance behaviour | 0.093<br>(-0.195 - 0.382) |
| Pre-intervention: Likely to get HIV vaccine [1-10] | 0.028<br>(-0.028 - 0.085) |
| Observations | 2,163 |
| Reference-group mean | 0.105 |

95% CI in parentheses
*** p<0.01, ** p<0.05
Outcome: Attrition in Follow-up Survey

**Table S7. OLS Estimates of the Intervention Effect on Self-Reported Credibility, Anxiety, and Willingness to Share Misinformation Claims Among Women Aged 18–29 in South Africa in the 3-week Follow-up Survey**

| | (1) | (2) | (3) | (4) | (5) | (6) | (7) | (8) |
|---|---|---|---|---|---|---|---|---|
| | Claim 1 | Claim 2 | Claim 8 | Claim 9 | Claim 10 | Claim 11 | Claim 12 | Claim 13^ |
| | | | | Credibility (1-7 scale) | | | | |
| Intervention | -0.930*** | -1.619*** | -0.578*** | -1.202*** | -0.387*** | -0.542*** | -0.461*** | -0.289*** |
| | (-1.047 - -0.812) | (-1.785 - -1.453) | (-0.713 - -0.443) | (-1.378 - -1.026) | (-0.476 - -0.298) | (-0.651 - -0.434) | (-0.581 - -0.340) | (-0.465 - -0.114) |
| Covariates | Yes | Yes | Yes | Yes | Yes | Yes | Yes | Yes |
| Observations | 1,936 | 1,936 | 1,936 | 1,936 | 1,936 | 1,936 | 1,936 | 1,936 |
| R-squared | 0.148 | 0.177 | 0.052 | 0.099 | 0.068 | 0.082 | 0.055 | 0.015 |
| Control-group mean | 2.327 | 3.736 | 2.142 | 3.918 | 1.581 | 1.918 | 2 | 5.022 |
| | | | | Anxiousness (1-7 scale) | | | | |
| Intervention | -1.259*** | -1.010*** | -0.760*** | -0.963*** | -0.587*** | -0.731*** | -0.519*** | -0.379*** |
| | (-1.421 - -1.098) | (-1.156 - -0.864) | (-0.936 - -0.585) | (-1.119 - -0.807) | (-0.716 - -0.459) | (-0.871 - -0.592) | (-0.669 - -0.369) | (-0.569 - -0.188) |
| Covariates | Yes | Yes | Yes | Yes | Yes | Yes | Yes | Yes |
| Observations | | | | | | | | |
| R-squared | 1,936 | 1,936 | 1,936 | 1,936 | 1,936 | 1,936 | 1,936 | 1,936 |
| Control-group mean | 0.158 | 0.122 | 0.055 | 0.096 | 0.074 | 0.092 | 0.052 | 0.024 |
| | 3.145 | 2.899 | 2.798 | 2.967 | 2.021 | 2.339 | 2.359 | 4.528 |
| | | | | Shareability (1-7 scale) | | | | |
| Intervention | -0.792*** | -1.186*** | -0.618*** | -0.968*** | -0.394*** | -0.521*** | -0.520*** | -0.340*** |
| | (-0.920 - -0.663) | (-1.346 - -1.026) | (-0.759 - -0.477) | (-1.123 - -0.814) | (-0.497 - -0.291) | (-0.633 - -0.409) | (-0.650 - -0.391) | (-0.537 - -0.144) |
| Covariates | Yes | Yes | Yes | Yes | Yes | Yes | Yes | Yes |
| Observations | | | | | | | | |
| R-squared | 1,936 | 1,936 | 1,936 | 1,936 | 1,936 | 1,936 | 1,936 | 1,936 |
| Control-group mean | 0.111 | 0.129 | 0.060 | 0.100 | 0.066 | 0.076 | 0.078 | 0.026 |
| | 2.150 | 2.874 | 2.137 | 2.753 | 1.627 | 1.823 | 1.983 | 3.807 |

95% CI in parentheses
*** p<0.01, ** p<0.05
Outcomes: Credibility is measured on a scale of 1-7, where 1 corresponds to the lowest self-reported likelihood of believing the misinformation claim and 7 the highest. Anxiousness is measured on a scale of 1-7, where 1 corresponds to the lowest self-reported anxiety upon seeing the misinformation claim and 7 the highest. Shareability is measured on a scale of 1-7, where 1 corresponds to the lowest self-reported likelihood of sharing the misinformation claim and 7 the highest.
Covariates include demographic and information and health behaviour covariates. Demographic covariates include age, African/Black race, completion of 12th grade, residence in an informal settlement or township, Gauteng province residence, and any wage work in the past seven days. Information and health behaviour covariates include any household hunger in the past seven days, any depression, daily social media use, never having received a COVID-19 vaccine, ever having used PrEP, high perceived HIV risk, confidence in scientists (1–4 scale), and any information avoidance behaviour.
^Claim 13 is the ‘true’ claim that presented information considered legitimate.
Data on the shareability of Claim 7 was not collected due to a survey error.

**Table S8. OLS Estimates of the Intervention Effect on Post-Inoculation Vaccine Intentions Among Women Aged 18–29 in South Africa By Sample Characteristics in the Main Survey**

| | Age 25 or more | Prep use ever | Perceive HIV risk as high | Never received COVID-19 vaccine | High information avoidance |
|---|---|---|---|---|---|
| | (1) | (2) | (3) | (4) | (5) |
| Pre-inoculation vaccine intention | 0.665*** | 0.664*** | 0.664*** | 0.663*** | 0.664*** |
| | (0.637 - 0.693) | (0.636 - 0.692) | (0.636 - 0.692) | (0.635 - 0.692) | (0.636 - 0.692) |
| Intervention | 0.925*** | 0.961*** | 0.975*** | 0.878*** | 0.815*** |
| | (0.700 - 1.149) | (0.792 - 1.130) | (0.799 - 1.152) | (0.713 - 1.043) | (0.606 - 1.023) |
| Sub-group | -0.023 | 0.270** | 0.181* | -0.318*** | -0.517*** |
| | (-0.227 - 0.181) | (0.047 - 0.493) | (-0.029 - 0.391) | (-0.556 - -0.079) | (-0.724 - -0.311) |
| Intervention x Sub-group | -0.012 | -0.146 | -0.160 | 0.157 | 0.188 |
| | (-0.300 - 0.276) | (-0.453 - 0.161) | (-0.453 - 0.134) | (-0.169 - 0.482) | (-0.095 - 0.470) |
| Demographic covariates | Yes | Yes | Yes | Yes | Yes |
| Information and health behaviours covariates | Yes | Yes | Yes | Yes | Yes |
| Observations | 2,163 | 2,160 | 2,163 | 2,127 | 2,163 |
| R-squared | 0.629 | 0.630 | 0.630 | 0.626 | 0.630 |
| Control-group mean | 7.178 | 7.178 | 7.178 | 7.178 | 7.178 |

95% CI in parentheses
*** p<0.01, ** p<0.05
Outcome: Post-inoculation main survey vaccine intention measured on a scale between 1-10, where 1 corresponds to the lowest intention of getting vaccinated and 10 the highest.
Demographic covariates include age, African/Black race, completion of 12th grade, residence in an informal settlement or township, Gauteng province residence, and any wage work in the past seven days. Information and health behaviour covariates include any household hunger in the past seven days, any depression, daily social media use, never having received a COVID-19 vaccine, ever having used PrEP, high perceived HIV risk, confidence in scientists (1–4 scale), and any information avoidance behaviour.
For the HIV PrEP variable, responses noting “Prefer not to say” (n=3) are excluded. For the COVID-19 vaccination variable, responses noting “Prefer not to say” (n=15) and “Don’t remember” (n=21) are excluded.

**Table S9. OLS Estimates of the Intervention Effect on Post-Inoculation Vaccine Intentions Among Women Aged 18–29 in South Africa By Sample Characteristics in the 3-week Follow-up Survey**

| | Age 25 or more | Prep use ever | Perceive HIV risk as high | Never received COVID-19 vaccine | High information avoidance |
|---|---|---|---|---|---|
| | (1) | (2) | (3) | (4) | (5) |
| Pre-inoculation vaccine intention | 0.464*** | 0.463*** | 0.463*** | 0.459*** | 0.463*** |
| | (0.425 - 0.502) | (0.424 - 0.502) | (0.424 - 0.502) | (0.420 - 0.498) | (0.424 - 0.501) |
| Intervention | 1.055*** | 1.035*** | 1.142*** | 0.771*** | 0.866*** |
| | (0.744 - 1.366) | (0.802 - 1.268) | (0.898 - 1.386) | (0.545 - 0.998) | (0.577 - 1.156) |
| Sub-group | 0.204 | 0.548*** | 0.347** | -0.929*** | -0.745*** |
| | (-0.078 - 0.486) | (0.237 - 0.860) | (0.058 - 0.636) | (-1.256 - -0.601) | (-1.031 - -0.458) |
| Intervention x Sub-group | -0.144 | -0.221 | -0.474** | 0.834*** | 0.189 |
| | (-0.543 - 0.254) | (-0.648 - 0.206) | (-0.880 - -0.068) | (0.388 - 1.280) | (-0.202 - 0.580) |
| Demographic covariates | Yes | Yes | Yes | Yes | Yes |
| Information and health behaviours covariates | Yes | Yes | Yes | Yes | Yes |
| Observations | 1,936 | 1,933 | 1,936 | 1,907 | 1,936 |
| R-squared | 0.424 | 0.423 | 0.425 | 0.426 | 0.423 |
| Control-group mean | 6.818 | 6.818 | 6.818 | 6.818 | 6.818 |

95% CI in parentheses
*** p<0.01, ** p<0.05
Outcome: Post-inoculation follow-up survey vaccine intention measured on a scale between 1-10, where 1 corresponds to the lowest intention of getting vaccinated and 10 the highest.
Demographic covariates include age, African/Black race, completion of 12th grade, residence in an informal settlement or township, Gauteng province residence, and any wage work in the past seven days. Information and health behaviour covariates include any household hunger in the past seven days, any depression, daily social media use, never having received a COVID-19 vaccine, ever having used PrEP, high perceived HIV risk, confidence in scientists (1–4 scale), and any information avoidance behaviour.
For the HIV PrEP variable, responses noting "Prefer not to say" (n=3) are excluded. For the COVID-19 vaccination variable, responses noting "Prefer not to say" (n=12) and "Don't remember" (n=17) are excluded.

**Table S10. OLS Estimates of the Intervention Effect on Post-Inoculation Vaccine Recommendations Among Women Aged 18–29 in South Africa By Sample Characteristics in the Main Survey**

| | Age 25 or more | Prep use ever | Perceive HIV risk as high | Never received COVID-19 vaccine | High information avoidance |
|---|---|---|---|---|---|
| | (1) | (2) | (3) | (4) | (5) |
| Pre-inoculation vaccine recommendation | 0.639*** | 0.637*** | 0.637*** | 0.637*** | 0.636*** |
| | (0.606 - 0.672) | (0.604 - 0.670) | (0.604 - 0.670) | (0.604 - 0.670) | (0.603 - 0.669) |
| Intervention | 0.691*** | 0.802*** | 0.767*** | 0.593*** | 0.492*** |
| | (0.475 - 0.907) | (0.640 - 0.964) | (0.598 - 0.937) | (0.436 - 0.751) | (0.291 - 0.692) |
| Sub-group | -0.091 | 0.189* | 0.307*** | -0.302*** | -0.748*** |
| | (-0.287 - 0.105) | (-0.025 - 0.402) | (0.106 - 0.508) | (-0.529 - -0.075) | (-0.948 - -0.549) |
| Intervention x Sub-group | 0.081 | -0.199 | -0.075 | 0.563*** | 0.456*** |
| | (-0.196 - 0.358) | (-0.494 - 0.096) | (-0.357 - 0.207) | (0.253 - 0.873) | (0.185 - 0.726) |
| Demographic covariates | Yes | Yes | Yes | Yes | Yes |
| Information and health behaviours covariates | Yes | Yes | Yes | Yes | Yes |
| Observations | 2,163 | 2,160 | 2,163 | 2,127 | 2,163 |
| R-squared | 0.542 | 0.543 | 0.543 | 0.547 | 0.545 |
| Control-group mean | 7.892 | 7.892 | 7.892 | 7.892 | 7.892 |

95% CI in parentheses
*** p<0.01, ** p<0.05
Outcome: Post-inoculation main survey vaccine recommendation measured on a scale between 1-10, where 1 corresponds to the lowest likelihood of recommending the vaccine to friends and 10 the highest.
For the HIV PrEP variable, responses noting “Prefer not to say” (n=3) are excluded. For the COVID-19 vaccination variable, responses noting “Prefer not to say” (n=15) and “Don’t remember” (n=21) are excluded.
Demographic covariates include age, African/Black race, completion of 12th grade, residence in an informal settlement or township, Gauteng province residence, and any wage work in the past seven days. Information and health behaviour covariates include any household hunger in the past seven days, any depression, daily social media use, never having received a COVID-19 vaccine, ever having used PrEP, high perceived HIV risk, confidence in scientists (1–4 scale), and any information avoidance behaviour.

**Table S11. OLS Estimates of the Intervention Effect on Post-Inoculation Vaccine Recommendations Among Women Aged 18–29 in South Africa By Sample Characteristics in the 3-week Follow-up Survey**

| | Age 25 or more | Prep use ever | Perceive HIV risk as high | Never received COVID-19 vaccine | High information avoidance |
|---|---|---|---|---|---|
| | (1) | (2) | (3) | (4) | (5) |
| Pre-inoculation vaccine recommendation | 0.483*** | 0.481*** | 0.481*** | 0.477*** | 0.481*** |
| | (0.436 - 0.529) | (0.434 - 0.527) | (0.434 - 0.527) | (0.430 - 0.523) | (0.434 - 0.527) |
| Intervention | 0.900*** | 0.957*** | 0.948*** | 0.651*** | 0.802*** |
| | (0.591 - 1.208) | (0.726 - 1.187) | (0.706 - 1.190) | (0.427 - 0.876) | (0.515 - 1.089) |
| Sub-group | -0.009 | 0.649*** | 0.262* | -0.678*** | -0.689*** |
| | (-0.289 - 0.270) | (0.341 - 0.957) | (-0.023 - 0.547) | (-1.001 - -0.354) | (-0.975 - -0.403) |
| Intervention x Sub-group | -0.108 | -0.417* | -0.319 | 0.753*** | 0.057 |
| | (-0.503 - 0.287) | (-0.839 - 0.006) | (-0.721 - 0.084) | (0.311 - 1.194) | (-0.331 - 0.444) |
| Demographic covariates | Yes | Yes | Yes | Yes | Yes |
| Information and health behaviours covariates | Yes | Yes | Yes | Yes | Yes |
| Observations | 1,936 | 1,933 | 1,936 | 1,907 | 1,936 |
| R-squared | 0.366 | 0.368 | 0.367 | 0.371 | 0.367 |
| Control-group mean | 7.212 | 7.212 | 7.212 | 7.212 | 7.212 |

95% CI in parentheses
*** p<0.01, ** p<0.05
Outcome: Post-inoculation follow-up survey vaccine recommendation measured on a scale between 1-10, where 1 corresponds to the lowest likelihood of recommending the vaccine to friends and 10 the highest. Demographic covariates include age, African/Black race, completion of 12th grade, residence in an informal settlement or township, Gauteng province residence, and any wage work in the past seven days. Information and health behaviour covariates include any household hunger in the past seven days, any depression, daily social media use, never having received a COVID-19 vaccine, ever having used PrEP, high perceived HIV risk, confidence in scientists (1–4 scale), and any information avoidance behaviour.
For the HIV PrEP variable, responses noting “Prefer not to say” (n=3) are excluded. For the COVID-19 vaccination variable, responses noting “Prefer not to say” (n=12) and “Don’t remember” (n=17) are excluded.

**Table S12. OLS Estimates of the Intervention Effect on Post-Inoculation Vaccine Intentions and Recommendations Among Women Aged 18–29 in South Africa by residence in an informal settle/township**

| Outcome: | Vaccine Intention | Vaccine Recommendation |
|---|---|---|
| VARIABLES | (1) | (2) |
| Pre-inoculation vaccine intention | 0.664*** | 0.637*** |
| | (0.636 - 0.692) | (0.604 - 0.670) |
| Intervention | 0.902*** | 0.845*** |
| | (0.727 - 1.077) | (0.677 - 1.013) |
| Sub-group | -0.058 | 0.212** |
| | (-0.266 - 0.149) | (0.013 - 0.411) |
| Intervention x Sub-group | 0.042 | -0.296** |
| | (-0.253 - 0.337) | (-0.579 - -0.013) |
| Demographic covariates | Yes | Yes |
| Information and health behaviours covariates | Yes | Yes |
| Observations | 2,163 | 2,163 |
| R-squared | 0.629 | 0.543 |
| Control-group mean | 7.178 | 7.892 |

95% CI in parentheses
*** p<0.01, ** p<0.05
Outcomes:Post-inoculation follow-up survey vaccine intention measured on a scale between 1-10, where 1 corresponds to the lowest intention of getting vaccinated and 10 the highest. Post-inoculation follow-up survey vaccine recommendation measured on a scale between 1-10, where 1 corresponds to the lowest likelihood of recommending the vaccine to friends and 10 the highest.
Demographic covariates include age, African/Black race, completion of 12th grade, residence in an informal settlement or township, Gauteng province residence, and any wage work in the past seven days. Information and health behaviour covariates include any household hunger in the past seven days, any depression, daily social media use, never having received a COVID-19 vaccine, ever having used PrEP, high perceived HIV risk, confidence in scientists (1–4 scale), and any information avoidance behaviour.

#### Supplementary Figures

**Figure S1**. **Study procedures and timing of collection of study outcomes**

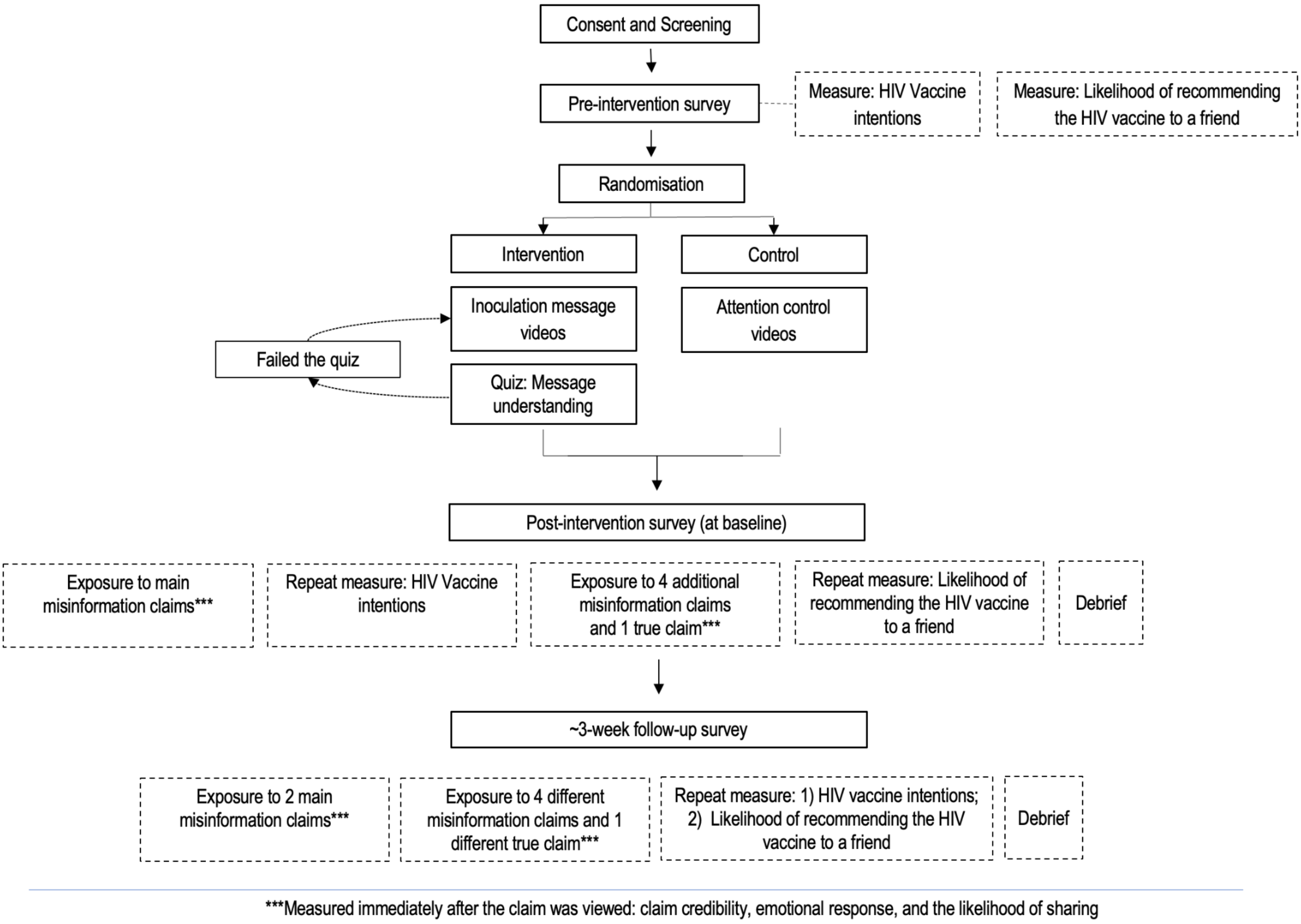

**Figure S2: Mean likelihood of getting the HIV vaccine across survey rounds**

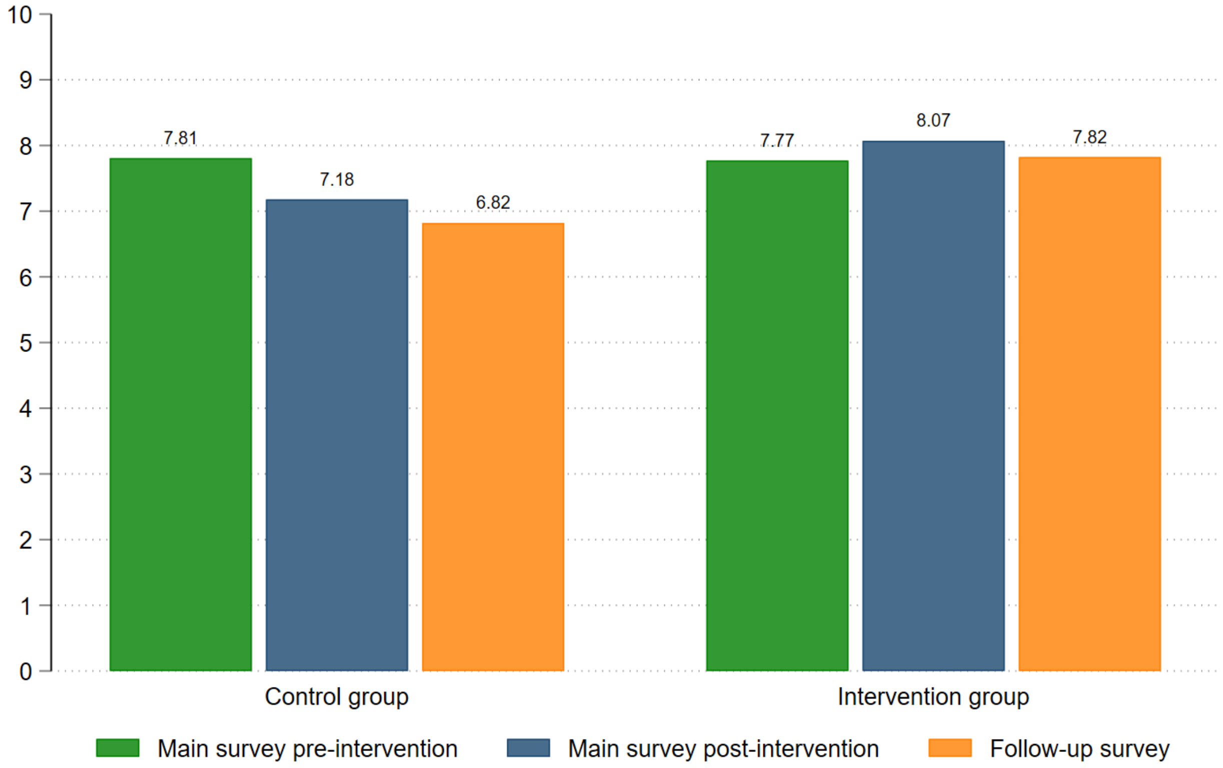


The primary outcome was the respondents' intention to get vaccinated. Specifically, participants were asked: "Now on a scale from 1 to 10, where 1 is extremely unlikely and 10 is extremely likely, please indicate how you feel about the following statements about the HIV vaccine: If an HIV vaccine became available, how likely would you be to get vaccinated?"

**Figure S3: Mean Likelihood of Recommending the HIV Vaccine across Survey Rounds**

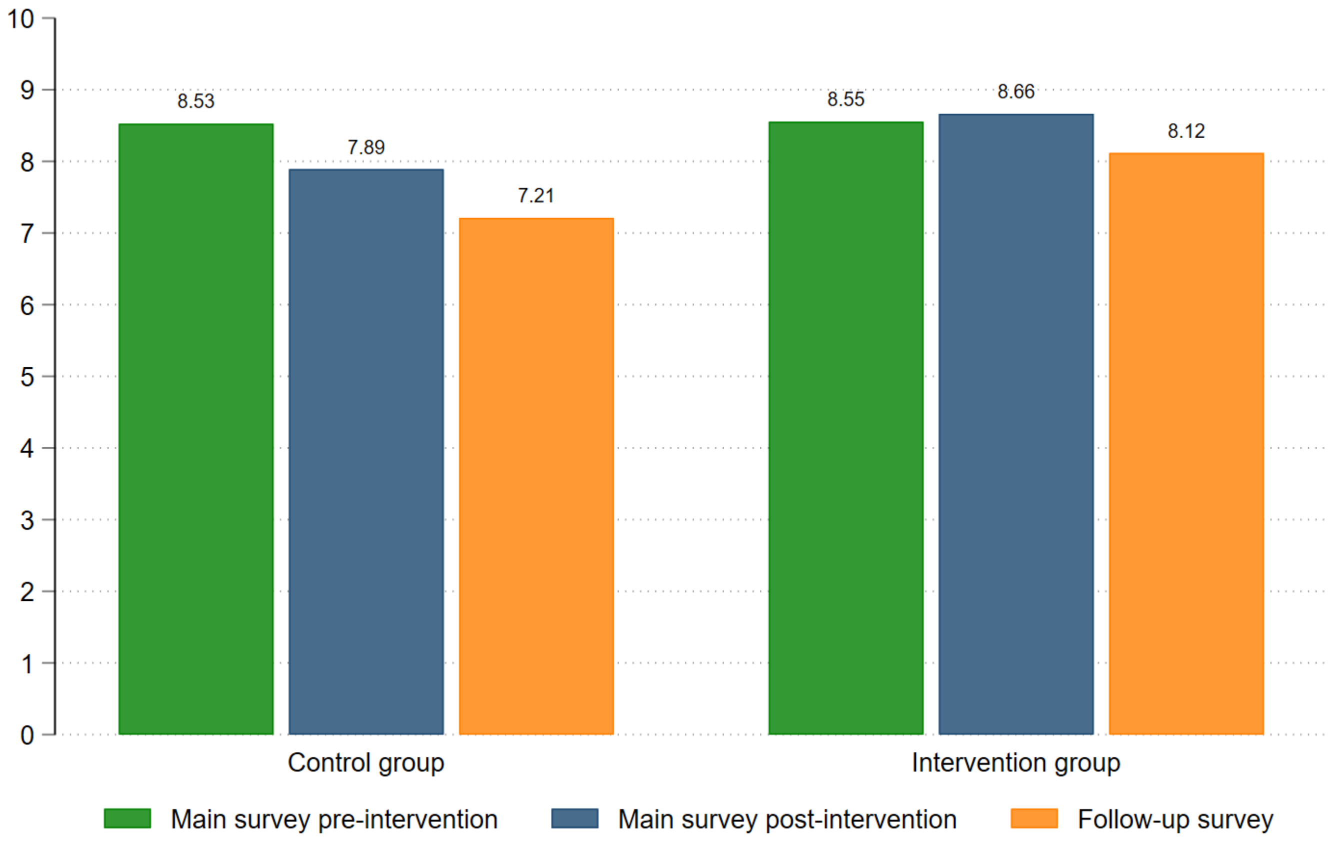


This secondary outcome assessed participants' likelihood of recommending the vaccine to friends: "On a scale from 1 to 10, where 1 is extremely unlikely and 10 is extremely likely, please indicate how you feel about the following statement:  If an HIV vaccine became available, how likely would you be to recommend it to your friends?"

**Supplementary Appendix 2**: PROTECT Inoculation Video Quiz

**Article**: Psychological inoculation protects young women in South Africa against HIV prevention misinformation: Evidence from a randomised controlled trial

**Video 1: Causal misattribution, or the *post hoc f*allacy**

**Introduction for the Quizzes**
Next, you'll watch two short videos about the HIV vaccine. We will also share some cool tricks with you along the way for how to spot fake information. Think about this:

Have you ever come across something online and wondered, Is this even true? Don't worry—we've got you covered! In this study, we'll give you some super useful tips to help you spot misinformation and avoid being misled. It's all packed into these two short videos, and we promise they're worth your time!

Please note you can only continue to the next page, once you have finished watching the video

**Framing for the quiz**
Thanks for watching the video! Here's a few quick questions to see whether you picked up the awesome tips for spotting and avoiding misinformation.

**Framing after getting the quiz correct and moving on to video 2**
You've totally got tip #1 for spotting and avoiding misinformation. Here's the next video with the second useful tip!

**Framing after getting the quiz wrong and being asked to watch video 1 again**
Thank you for watching the video and answering the questions! We noticed that a few answers were incorrect, and that's totally okay, it's all part of the learning process. Tip 1 for spotting and avoiding misinformation is super useful so we're going to show you the video again to give you the chance to hear this tip once more.

**Rule for who watches the video again:**
- Any incorrect answers result in them being asked to watch the video again.

**Quiz**

Your favorite TikToker says she got asked out by her crush because she got a new haircut. Why else could she have been asked on a date?
a. She's fun to be around
b. Her crush finally worked up the confidence to ask
c. Their mutual friend suggested that they go out
d. All of the above

    - For any response, the following text appears: “Well done. You are correct: Two things can happen one after the other but have nothing to do with each other. There are many other reasons why she might have been asked out”

In summer, ice cream sales increase and so do shark attacks. Do you think that ice cream sales cause shark attacks?

- Response options: Yes or No or Don’t Know
- If “Yes” or “Don’t Know” the following text appears: “Nice try, but this is not possible. Remember: Two things can happen at the same time and have nothing to do with each other..”
- If “No” the following text appears: “Well done. You are correct: Two things can happen at the same time and have nothing to do with each other.”

Your friend tells you she got the flu vaccine and then two weeks later she was diagnosed with cancer. Do you believe that her getting the vaccine caused her to get cancer?

- Response options: Yes or No or Don’t Know
- If “Yes” or “Don’t Know” the following text appears: “Nice try, but this is not how cancer works. Remember: Two things can happen one after the other but have nothing to do with each other.”
- If “No” the following text appears: “Well done. You are correct: Two things can happen one after the other but have nothing to do with each other.”

#### <u>Video 2: Impossible expectations</u>

**Framing after getting the quiz correct and moving on to the survey**
You've totally got tip #2 for spotting and avoiding misinformation. Time for the next section!

**Framing after getting the quiz wrong and being asked to watch video 2 again**
Thank you for watching the video and answering the questions! We noticed that a few answers were incorrect, and that's totally okay, it's all part of the learning process. Tip 2 for spotting and avoiding misinformation is super useful so we're going to show you the video again to give you the chance to hear this tip once more.

**Video 2 Quiz**

Which of the following has ZERO risk?

a. Driving in a car
b. Going for a walk
c. Taking a panado
d. None of the above

- If "a", the following text appears: "Nice try, but remember it is IMPOSSIBLE for this to have ZERO risk. We do this even though it is not 100% safe.."
- If "b", the following text appears: "Nice try, but remember it is IMPOSSIBLE for this to have ZERO risk. We do this even though it is not 100% safe."
- If "c", the following text appears: "Nice try, but remember it is IMPOSSIBLE for this to have ZERO risk. We do this even though it is not 100% safe."
- If "d", the following text appears: "Well done! This is correct. It is IMPOSSIBLE for these activities to have ZERO risk. We do them even though it is not 100% safe.."

Do you think this statement is misleading: "People should not use birth control because it is not 100% safe"

a. Yes, it is misleading
b. No, it is not misleading

- If "a", the following text appears: "Well done! You're correct! It is IMPOSSIBLE for this to have ZERO risk. People take birth control pills even though there is a small chance of side effects or health risks."
- If "b", the following text appears: "Nice try, but remember it is IMPOSSIBLE for this to have ZERO risk. People take birth control pills even though there is a small chance of side effects or health risks"

You need to fly from Joburg to Cape Town but someone tells you not to go because airplanes are not 100% safe. Do you think this is a good reason not to go?

- Response options: Yes or No or Don’t Know
- If “Yes” or “Don’t know”, the following text appears: “Nice try, but remember: It is IMPOSSIBLE for travel to have ZERO risk. If you only travel when there is zero risk, you would never go anywhere. Zero risk is an impossible expectation - it’s not how the world works.
- If “No” the following text appears: “Well done. You are correct. It is IMPOSSIBLE for travel to have ZERO risk. If you only travel when there is zero risk, you would never go anywhere. Zero risk is an impossible expectation - it’s not how the world works.”

**Supplementary Appendix 3**: PROTECT Facebook Post Claims

**Article**: Psychological inoculation protects young women in South Africa against HIV prevention misinformation: Evidence from a randomised controlled trial

#### Baseline survey

(Fake) Facebook username associated with all claims: Slay Queen

##### Main misinformation claims

1. My sister got the HIV vaccine and her heart stopped the next day. Please don't get vaccinated- IT COULD KILL YOU 😭
2. HIV vaccine?! Yea, right, as soon as you can prove it's ZERO risk! #HIVvaccine #nomorejabs

##### Additional misinformation claims

3. The HIV vaccine gave my best friend HIV
4. The HIV vaccine isn't worth getting unless it's 100% effective in preventing HIV
5. That COVID vaccine? I think the government made it so African women couldn't have children
6. Vaccines! No way! We should not be getting these until we get a 100% promise that there are absolutely no side-effects.

##### 'True' claim

7. Ok, this HIV vaccine sounds cool, hey? I like that they will only start giving it once it's tested and approved 👍.

#### Follow-up survey

##### Main misinformation claims

1. My sister got the HIV vaccine and her heart stopped the next day. Please don't get vaccinated- IT COULD KILL YOU 😭
2. HIV vaccine?! Yea, right, as soon as you can prove it's ZERO risk! #HIVvaccine #nomorejabs

##### Additional misinformation claims

8. 🪰 👂 WTFly!?! A fly landed on my aunt's ear and the next day she went deaf in that ear!
9. I got the flu vaccine last year and still got flu, hello?!? Why take a vaccine that's not 100% effective?
10. The HIV vaccine goes inside your body and changes your blood into virus. You're an HIV Zombie!
11. Taking PrEP pills every day stops your ovaries from working properly and then you can't get pregnant
12. Dr. Matthew Lani says Vitamin C prevents cancer! Take Vitamin C for no cancer!

##### 'True' claim

13. Smoking literally increases your risk for lung cancer for sure -- that's been proven! But that doesn't mean everyone who smokes gets lung cancer
